# Augmenting vaccine efficacy against delta variant with ‘Mycobacterium-*w*’ mediated modulation of NK-ADCC and TLR-MYD88 pathways

**DOI:** 10.1101/2022.11.02.22281822

**Authors:** Sarita Rani Jaiswal, Ashraf Saifullah, Jaganath Arunachalam, Rohit Lakhchaura, Dhanir Tailor, Anupama Mehta, Gitali Bhagawati, Hemamalini Aiyer, Subhrajit Biswas, Bakulesh Khamar, Sanjay V. Malhotra, Suparno Chakrabarti

## Abstract

Mycobacterium-w (Mw), was shown to boost adaptive natural killer (ANK) cells and protect against COVID-19 during the first wave of the pandemic. As a follow-up of the trial, 50 healthcare workers (HCW) who had received Mw in September 2020 and subsequently received at least one dose of ChAdOx1 nCoV-19 vaccine (Mw+ChAdOx1 group) were monitored for symptomatic COVID-19, during a major outbreak with the delta variant of SARS-CoV-2 (April-June, 2021), along with 201 HCW receiving both doses of the vaccine without Mw (ChAdOx1 group). Despite 48% having received just a single dose of the vaccine in Mw+ChAdOx1 group, only 2 had mild COVID-19, compared to 36 infections in the ChAdOx1 group (HR-0.46, p=0.009). Transcriptomic studies revealed an enhanced adaptive NK cell-dependent ADCC in the Mw+ChAdOx1 group, along with downregulation of TLR2-MYD88 pathway and concomitant attenuation of downstream inflammatory pathways. This might have resulted in robust protection during the pandemic with the delta variant.

## INTRODUCTION

SARS-CoV-2 pandemic has changed the prevailing concepts related to the timeline for development of novel vaccines. Collaborative efforts between scientists, industry and regulators resulted in effective vaccines for SARS-CoV-2 being available in less than 12 months. The vaccines used time tested technologies like inactivated virus as well as novel technologies like viral vector or mRNA to provide an antiviral response ^1^. While this feat remains a historical milestone, SARS-CoV-2 mutated at a rapid pace and has generated multiple variants of concerns (VOC) ^2,3^. VOCs are known to escape immune response generated by a previous infection/vaccination, leading to reduced/loss of protection against subsequent VOC as well as a decrease in vaccine efficacy ^4,5^. To improve the protection against VOC, booster doses of approved vaccines have been advocated periodically ^6,7^. This puts a significant burden on healthcare delivery system warranting a more robust solution.

Prior to availability of a vaccine during first wave of SARS-CoV-2 infection, our group, successfully explored the protection provided by the innate immune pathway, following its boosting using an approved Mycobacterium-*w* (Mw), an immunomodulator in high-risk health care workers (HCW) ^8^. Protection provided following Mw administration, in the absence of any specific agents for prevention or treatment, was associated with the upregulation of adaptive natural killer (ANK) cells and Antibody-dependent cellular cytotoxicity (ADCC) pathway, which was shown to provide protection against symptomatic COVID-19.

In the second quarter of 2021, India witnessed an unprecedented outbreak with the delta variant (B.1.617 lineage) of SARS-CoV-2, which despite the introduction of two effective vaccines against SARS-CoV-2 (ChAdOx1 nCoV-19 and BBV152) by February 2021, overwhelmed the system with its rapid transmission and fatality, not quite witnessed during the first wave of the pandemic ^9^. By this time (February 2021) we had evaluated our clinical trial data using Mw for protect against COVID -19 and were following up the cohort for long term outcome. We took this opportunity to evaluate the impact of Mw administration on protection mediated by ChAdOx1 nCoV-19 vaccine in a cohort-control study and understand the mechanistic pathways behind this by transcriptome analysis in the two groups.

## RESULTS

The current study reports the outcome of Mw + ChAdOx1 group vs ChAdOx1 group in terms of COVID-19, along with the gene expression profile, with regard to inflammatory as well as ANK-ADCC pathways.

### Effect of Mw priming on incidence and outcome of COVID-19

Both groups were identical in relation to age and gender (table 1). However, infection during first wave of COVID-19, prior to receiving of vaccine was significantly higher in the ChAdOx1 group. Similarly, all in the ChAdOx1 group had received both doses of the vaccine, but only 24/50 (48%) subjects in Mw + ChAdOx1 group had received both doses (p=0.0001).

During the study period, thirty-eight out of 251 (15.1%) subjects developed symptomatic COVID-19 infections. Symptomatic COVID-19 infection rate was significantly lower (4%) in the Mw + ChAdOx1 group compared to 17.9% in the ChAdOx1 group (p=0.01) (Figure 1).

**Figure 1:**
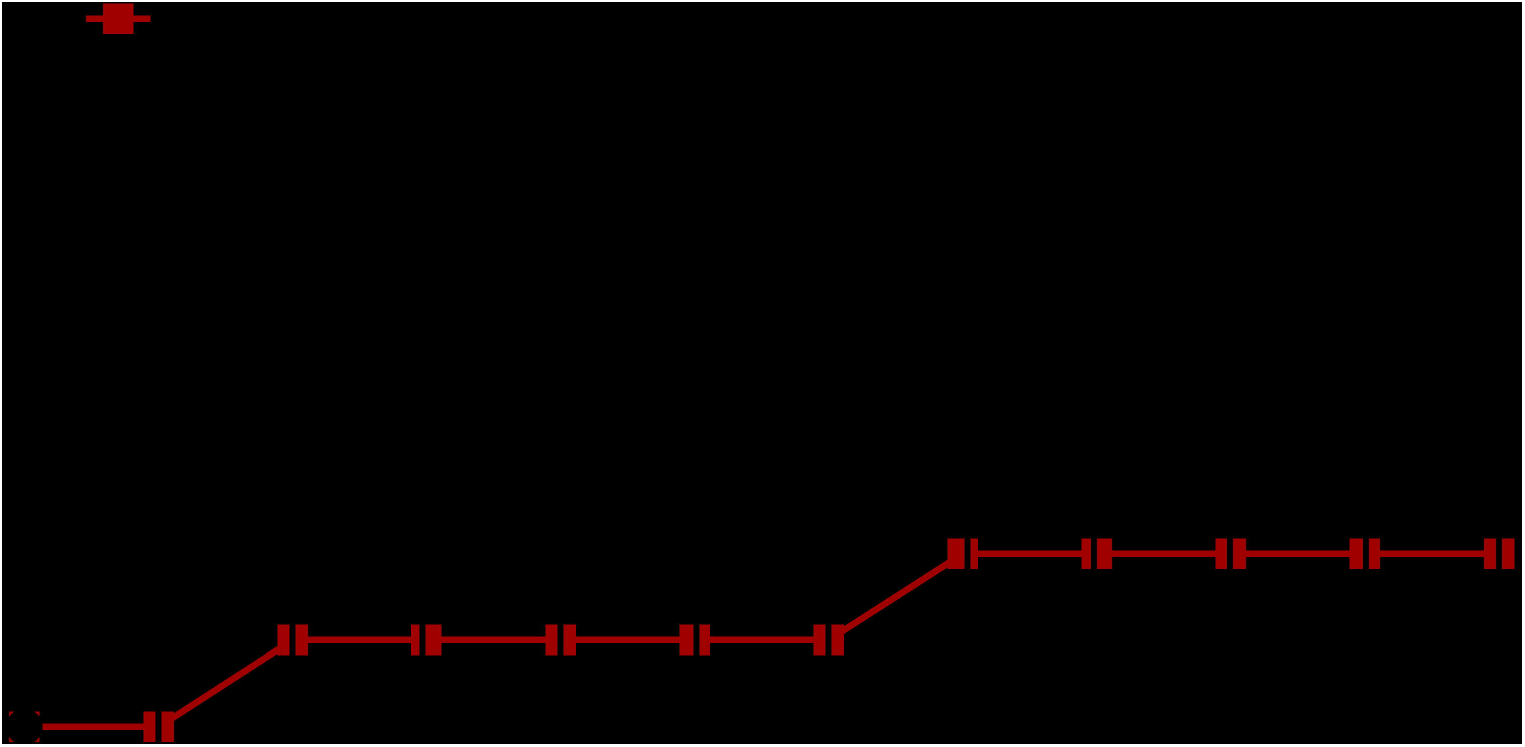
Impact of Mw vaccination on COVID-19 compared to ChAdOx1 group: Points and connecting line plot show the infection trend in Mw + ChAdOx1 group (n=50) and the ChAdOx1 group (n=201). The x-axis shows the time in weeks and y-axis shows the cumulative incidence (CI) in %. Red and black shaded circles represent Mw + ChAdOx1 and ChAdOx1 groups respectively.

The incidence rate (IR) of COVID-19 in the Mw + ChAdOx1 group was 6.7 infections/10,000 person-days, compared to 29.85 infections/10,000 person-days in the ChAdOx1 group, with an incidence rate ratio of 0.22 (95% CI, 0.05-0.9, p=0.02) and hazard ratio (HR) of 0.46 (95% CI, 0.11-1.93, p=0.009, Supplement).

Severe COVID-19 requiring hospitalization was not seen in the Mw + ChAdOx1 group but was seen in 9 (25%) in the ChAdOx1 group. Reinfection rate was identical in both groups, 13 subjects in the ChAdOx1 group and one in the Mw + ChAdOx1 group (p=0.3). No deaths were recorded in either group.

### DGE Analysis

Sequencing and mapping metrics are explained in supplement table S1. Based on criteria of p-value ≤ 0.05 and a log2 fold change (log2FC) >0.5 & ≤-0.5, a total of 584 out of 14,148 genes were found to be differentially expressed between the two groups, with 223 and 361 genes significantly up and downregulated respectively in the Mw + ChAdOx1 group. Forty-six DEGs related to ANK-ADCC, and innate immune inflammatory pathways were selected out to analyze the differential expression pattern between two groups (Figure 2A and 2B).

**Figure 2:**
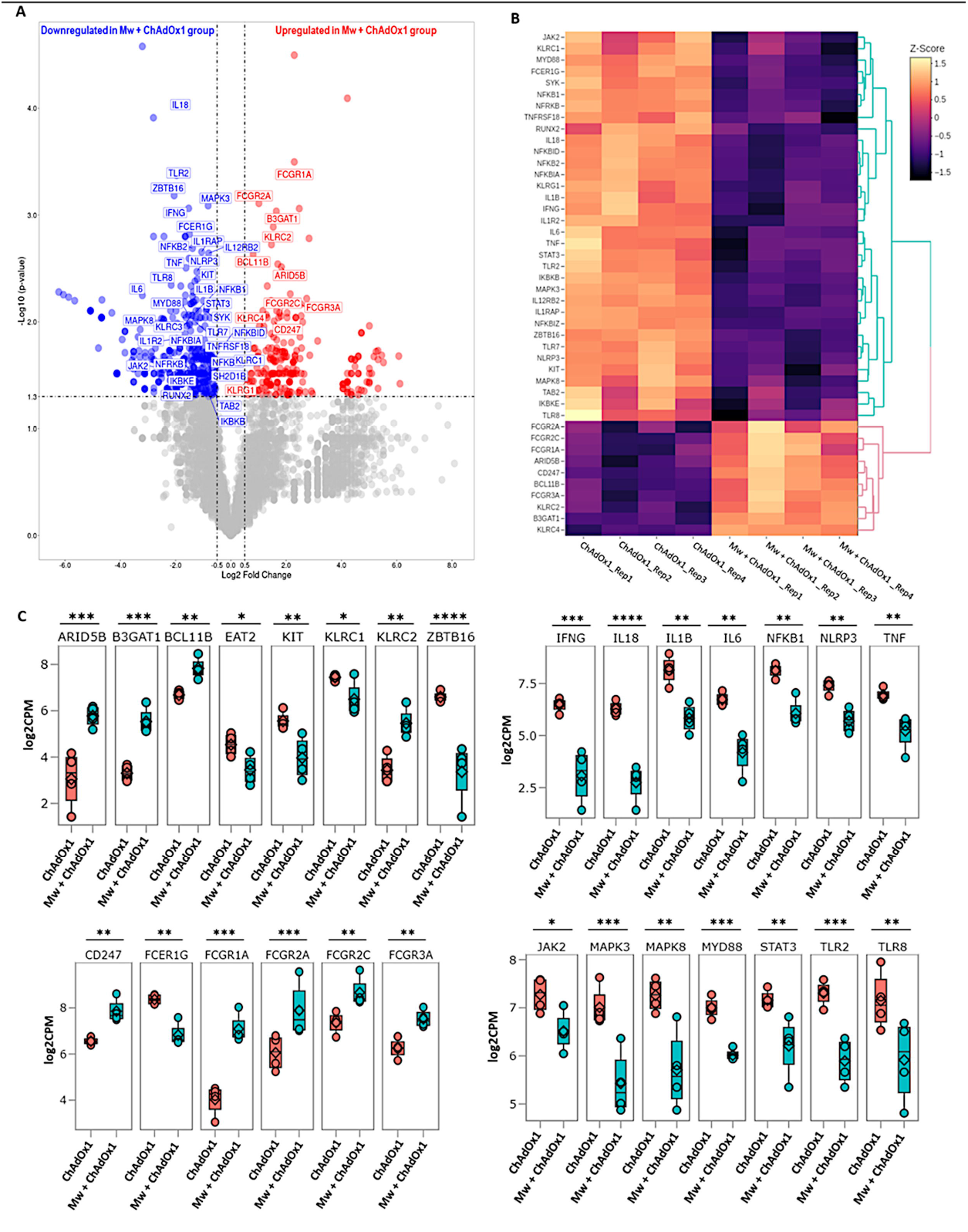
Differential gene expression analysis in Mw + ChAdOx1 and ChAdOx1 group: A) Volcano plot showing genes differentially expressed in Mw + ChAdOx1 as compared to ChAdOx1 group. Genes with a p-value <0.05 and log2 Fold Change ≥0.5 & ≤-0.5 were considered significant. Selected genes are highlighted (Red: Upregulated in Mw + ChAdOx1 group, Blue: Downregulated in Mw + ChAdOx1 group). B) Heatmap of RNA-Seq expression data showing selected DEGs related to this study. Hierarchical cluster analysis was performed between all samples of Mw + ChAdOx1 and ChAdOx1 group employing normalized log2 counts per million (log2CPM) expression data. The colour gradient scale indicates z-score C) Boxplots showing expression levels of 28/46 selected genes in Mw _+_ ChAdOx1 (green) as compared to ChAdOx1 group (orange). Relative gene expression is shown in normalized log2CPM for I) ANK-ADCC pathway. II) Innate immune inflammatory pathway, (*p-value < 0.05, **p-value < 0.01, ***p-value < 0.001, ****p-value<0.0001).

### ANK-ADCC pathway genes

Amongst 19 DEGs associated with ANK and ADCC pathways, 11 genes were upregulated and 8 were downregulated in Mw + ChAdOx1 group. KLRC2 (NKG2C), BCL11B, ARID5B, B3GAT1 (CD57), KLRC4 were upregulated and KLRC1 (NKG2A), ZBTB16 (PLZF), KIT and SH2D1B (EAT-2) were downregulated in relation to ANK pathway. In the ADCC pathway, CD247 (CD3ζ), FCGR1A (CD64), FCGR2A (CD32a), FCGR2C (CD32c) and Fc Gamma Receptor IIIa (FCγRIIIA, CD16a) were upregulated and FCER1G was downregulated (Figure 2C). This profile was characteristic of an upregulated ANK mediated ADCC pathway.

### Innate Immune Inflammatory pathway genes

Among 27 DEGs associated with innate inflammatory pathways Interferon Gamma (IFNγ), Tumor Necrosis Factor-Alpha (TNFα), Interleukin 1 Beta (IL1β), Interleukin-6 (IL6), Interleukin-8 (IL18), Janus Kinase 2 (JAK2), Signal Transducer And Activator Of Transcription 3 (STAT3), Myeloid Differentiation Primary Response 88 (MYD88), Toll-Like Receptor-2 (TLR2), Toll-Like Receptor-7 (TLR7), Nuclear Factor Kappa B Subunit-1 (NF-κB1), NOD Like Receptor Family Pyrin Domain Containing 3 (NLRP3), Mitogen-Activated Protein Kinase-3 (MAPK3) and Mitogen-Activated Protein Kinase-8 (MAPK8) were found to be significantly downregulated in Mw + ChAdOx1 group (Figure 2C).

### GO Pathway Analysis

The pathway enrichment of selected DEGs was carried out focusing on ANK-ADCC and innate immune inflammatory pathways using ClusterProfiler package employing GO database. Among significant pathways of ANK-ADCC pathway, antibody-dependent cellular cytotoxicity (GO:0001788); positive regulation of natural killer cell mediated cytotoxicity (GO:0045954) and innate immune response activating cell surface receptor signalling pathway (GO:0002220) were enriched for upregulation. Thirteen major innate inflammatory associated pathways were enriched for downregulation in the Mw + ChAdOx1 group. These included MyD88-dependent toll-like (GO:0002755), positive regulation of toll-like receptor (GO:000224), receptor *via* JAK-STAT (GO:0046427) signaling pathway; positive regulation of cytokine production (GO:1900017), inflammatory response (GO:0050729); positive regulation of NIK/NF-kappaB signalling (GO:1901224); positive regulation of interleukin-1 beta (GO:0032731), interleukin-6 (GO:0032755), interleukin-8 (GO:0032757), interleukin-12 (GO:0032735), interleukin-17 (GO:0032740) production; positive regulation of interferon-gamma (GO:0032729), tumor necrosis factor superfamily cytokine (GO:1903557) production (Figure 3).

**Figure 3:**
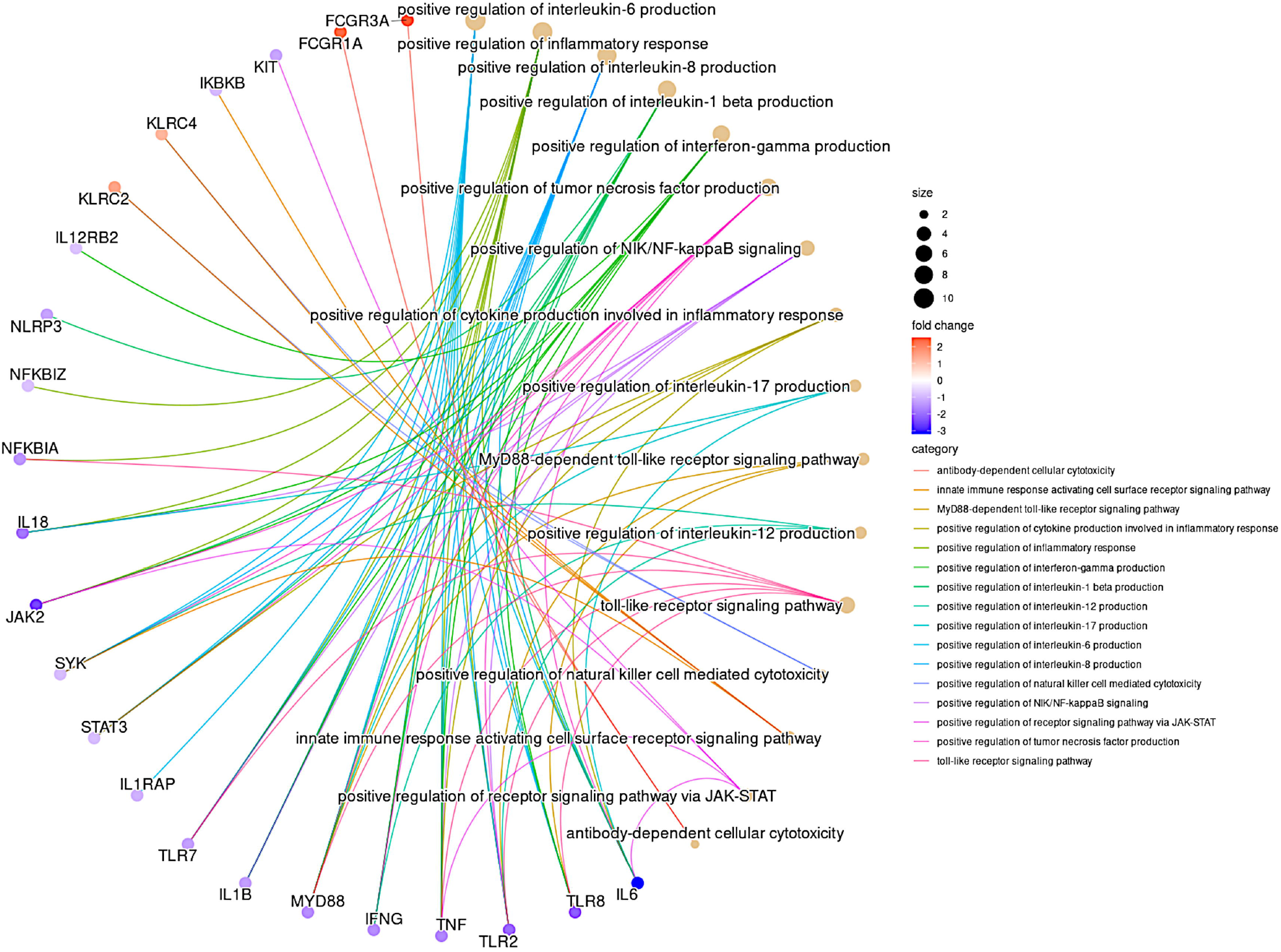
Gene Ontology pathway network analysis of DGEs: GO pathway network analysis of the top 16 enriched GO terms in the differentially expressed genes for ADCC and innate immune inflammatory pathway (Fisher’s exact test using enrichGO function in R package clusterProfiler, multiple test correction by Benjamini-Hochberg method, adj. p-value <0.05). The colour gradient scale indicates the log2FC of genes.

## DISCUSSION

During the study period of April-June 2021, India had witnessed a raging second wave of COVID-19, caused by the delta variant of SARS-CoV-2. Nineteen million infections were recorded in this period, with nearly one million cases in New Delhi, the national capital, where the study was undertaken ^10,11^. The efficacy of two doses of the ChAdOx1 nCoV-19 vaccine against the delta variant was subsequently reported to be 63.1% ^9^. Despite double dose vaccination, 17.9% of HCW in the control group were infected in our study, a quarter of them requiring oxygen support and hospitalization. This was similar to a nationwide survey, which reported a prevalence of breakthrough infections after two doses of vaccine in healthcare workers in India at 23.4% ^12^. In contrast, only 4% in the Mw + ChAdOx1 group got infected, with all experiencing mild infection.

Enhanced protection in the Mw + ChAdOx1 group during outbreak with delta variant was observed, despite only 48% receiving two doses of the vaccine in Mw + ChAdOx1 group, compared to all receiving both the doses in ChAdOx1 group. This suggests that factors other than neutralizing antibodies might be in play. ADCC is another known mechanism which operates through the engagement of cellular effectors *via* Fc receptors ^13^. Amongst these, NK cell mediated ADCC is the most potent and consistent. While VOC might escape neutralization by mutational changes in the receptor binding domains, NK cell mediated ADCC effector functions is more resilient and remain preserved across the variants ^14-17^ for longer periods ^18^.

We had earlier demonstrated that ANK cells might provide protection against SARS-CoV-2 and Mw could be providing protection *via* augmentation of ANK cells, an effect which seems to persist beyond 6 months ^8^. Both natural COVID-19 infection as well as the vaccines have been shown to produce potent NK cell mediated ADCC response ^15^. Several studies have shown that ADCC induced following vaccination is more resilient against mutants (VOC) in contrast to variable neutralizing antibody response against various VOC ^14,17^. Based on DEGs related to ANK-ADCC gene pathways in this study, ADCC generated following Mw priming of vaccination seems to be more potent compared to ChAdOx1 vaccine alone. Increased expression of CD247, along with downregulation of FC_ε_R1G observed in the Mw + ChAdOx1 group is a typical signature of ANK-ADCC ^19^. Both CD247 and FC_ε_R1G are adapter molecules for FCγRIIIA (CD16) with CD247 possessing three immunoreceptor Tyrosine-based activation motifs (ITAM) as against one ITAM of FCE°R1G, increasing ADCC by several folds ^20^. Thus, persistent upregulation of ANK cells and the augmented ADCC pathway via ANK cells following exposure to Mw might have boosted the protective efficacy of even a single dose of ChAdOx1 vaccine.

Disease severity in SARS-CoV-2 infection is a result of inappropriate hyperinflammatory cytokine response, resulting from increased release of IFNγ, TNFα, IL1β, IL6 and IL18, amongst others. In COVID-19, TLR2 has been shown to be the primary innate sensor of the envelope protein of SARS-CoV-2, generating a cytokine response mediated via the MYD88 pathway with subsequent activation of NF-κB and MAPK signaling ^21,22^. There was further downregulation of NLRP3-IL1β pathways, which have been also implicated in pathogenesis of COVID-19 ^23^. Based on the RNA-Seq findings, we hypothesize that Mw-mediated downregulation of the key TLR2-MYD88 pathway and its subsequent downstream signaling pathways, might have contributed substantially to its efficacy terms of absolute reduction in development of symptomatic COVID-19 as well as moderation of severity of COVID-19. Continued agonist-induced downregulation might be the most likely explanation for this phenomenon but would require longitudinal studies to confirm the same. Further limitations of our study were that it was carried out on a small cohort without randomization and lacked further exploration of other humoral, T cell and monocyte/macrophage mediated pathways at a cellular level. However, the distinct gene expression signature indicates a salutary effect of Mw priming on the NK-ADCC pathway and downregulation of the critical TLR2-MYD88 pathway at the same time, which might overcome limitations of vaccines against the current and evolving VOC.

## METHODS

The study was reviewed and approved by the institutional ethics committee. All subjects provided written consents for participation in the study. In this single-center non-randomized cohort control study, the effect of prior priming with Mw on the protection offered by ChAdOx1 nCoV-19 vaccine was evaluated during April to June 2021, at the peak of severe outbreak of delta variant of SARS-CoV2 in New Delhi, India. All enrolled subjects had their first dose of ChAdOx1 nCoV-19 vaccine (Covishield, Serum Institute of India, Pune, India) by February 28, 2021 of which 50 (Mw + ChAdOx1 group) had received 0.1 ml Mw (Sepsivac, Cadila Pharmaceuticals, India) intradermally in each arm in September, 2020 (≥ 6 months) as a part of a trial (CTRI/2020/10/028326) ^8^. Control arm included 201 HCWs from the same institute who received both doses of the ChAdOx1 nCoV-19 vaccine but not Mw, during or prior to their enrollment in the study (ChAdOx1 group). All subjects had their body temperature, pulse rate, oxygen saturation evaluated before and after each working day besides self-reporting of symptoms. Nasopharyngeal swab was taken on development of symptoms suggestive of COVID-19 or following contact with a patient or a family member diagnosed with COVID-19 for reverse transcriptase-polymerase chain reaction (RT-PCR) for diagnosis of COVID-19 ^24^ (supplement). The severity of COVID-19 was graded as per established criteria ^25^.

### Statistics

Binary variables were compared between the groups using chi square test. The continuous variables were analysed using independent sample t-test considering Levene’s test for equality of variances and non-parametric tests (Mann-Whitney U test). P value < 0.05 was considered to be significant.

The efficacy of Mw in reducing the incidence of COVID-19 was calculated in terms of attack rates incidence risk ratio (IRR), absolute risk reduction (ARR) and intervention efficacy (see Table S3). Hazard ratio was calculated by the Cox regression method (SPSSv24, Armonk, USA). GraphPad Prism (version 9.0 for Windows, GraphPad Software, La Jolla, CA, USA) was used for graphical representation of the data.

### RNA Sequencing (RNA-Seq) Analysis

This was carried out on both Mw + ChAdOx1 and ChAdOx1 groups at enrollment on 4 subjects from each group, as per established protocol ^8^ and is detailed in the supplement. In brief, RNA isolation from peripheral blood mononuclear cells was carried out using Trizol method and poly(A) RNA selection. Complementary DNA (cDNA) library preparation and whole transcriptome sequencing were undertaken using MinION 2.0 Oxford Nanopore Technologies (ONT).

DGE was done using “pipeline-transcriptome-de” (https://github.com/nanoporetech/pipeline-transcriptome-de) pipeline. This pipeline from nanopore tech uses snakemake, minimap2, salmon, edgeR, DEXSeq and stageR to automate DGE workflows on long read data. Pipeline was set to make only reads aligned to minimum 3 samples to be considered for analysis. A separate conda (https://docs.conda.io/en/latest/#) environment was created on Linux OS to host this pipeline. Quantification and DGE was done by R language based (https://www.r-project.org/) tool edgeR (https://bioconductor.org/packages/release/bioc/html/edgeR.html) employing gene-wise negative binomial regression model and normalisation factor (Transcript Mean of M-value) for each sequence library. Differentially expressed genes (DEGs) with log2Fold Change (log2Fc) ≥0.5, ≤-0.5 and associated p-value <0.05 were selected as significant for further analysis. Annotation of DEGs was fetched from ENSEMBL database (https://asia.ensembl.org/index.html) using R based biomaRt package (https://bioconductor.org/packages/release/bioc/html/biomaRt.html). All the file compilation was ultimately done using Microsoft Excel and Linux Libre Office calc.

Hierarchical clustering analyses was performed between all four samples of each group to generate Heatmap from normalised log2 counts per million (log2CPM) expression values using R/Shiny based rnaseqDRaMA (https://hssgenomics.shinyapps.io/RNAseq_DRaMA/) package. Comparative gene expression boxplot on the basis of log2CPM expression values between Mw + ChAdOx1 and ChAdOx1 group were generated from START (https://kcvi.shinyapps.io/START/) tool. Volcano plot involving all DEGs were created using R based ggplot2 (https://ggplot2.tidyverse.org/) package.

### Gene Ontology (GO) Pathway Analysis

Pathway enrichment analysis was done for significant DEGs using R based clusterProfiler (https://bioconductor.org/packages/release/bioc/html/clusterProfiler.html, v.4.2.1) tool employing Gene Ontology database. Pathways related to ADCC and innate immune inflammatory pathways were considered for focused analysis. An adjusted *p*-value threshold of ≤0.05 was considered for this study.

## Data Availability

All data produced in the present study are available in Gene Expression Omnibus(GEO) NCBI database with restricted access

## ACKNOWLEDGEMENTS

We thank all the nursing and technical staff of the department who have assisted in carrying out this study.

## AUTHOR CONTRIBUTIONS

SRJ, BK, and SC designed the study. AM, RL, GB, and HM performed the study. SRJ, AS, AM, GB, and HM collected the data. SJ, AS, JA, SM, DT and SC analyzed the data. SRJ and SC wrote the manuscript. All the co-authors reviewed and approved the manuscript.

