## Supplement file for "Augmenting vaccine efficacy against delta variant with ‘Mycobacterium-*w*’ mediated modulation of NK-ADCC and TLR-MYD88 pathways"

**Real‐time reverse transcription–polymerase chain (RT‐PCR) for detection of SARS‐CoV‐2**

All COVID tests were done by the TrueNat real‐time reverse transcription–polymerase chain reaction (RT‐PCR) test. Oropharyngeal or nasopharyngeal swabs were collected using a standard nylon‐flocked swab and inserted into the viral transport medium (VTM) provided by the same company (Molbio Diagnostics Pvt. Ltd., Goa, India). Samples were transported immediately to the laboratory maintaining proper temperature and processed as per the manufacturer’s guidelines (TrueNat Beta CoV chip‐based real‐time PCR test for betacoronavirus; Molbio Diagnostics Pvt. Ltd.). The target sequence for this assay is E gene of the Sarbeco virus and human RNase P (internal positive control). Confirmatory genes were RdRP gene and ORF1A gene.

**Methods for RNA Sequencing and Differential Gene Expression Analysis (DGE)**

**RNA isolation using Trizol method and poly(A) RNA selection**

Peripheral blood mononuclear cells (PBMC) were isolated from whole blood samples by density gradient centrifugations using HiSep™ LSM 1077 media (Himedia, Mumbai, India). 5 x 106 PBMCs were used for RNA isolation using the Trizol method and followed by DNase treatment for RNA purification. Purified RNA was quantified using a Qubit 4.0 fluorometer. Take 5μg of total RNA used for the poly(A) RNA selection using NEB NEXT oligo d(T)25 beads (NEB, MA, USA). Poly(A) RNA was quantified and integrity assessed using Qubit 4.0 fluorometer (Invitrogen, Waltham, Massachusetts, United States) according to manufacturer instructions.

**cDNA library preparation and whole transcriptome sequencing using MinION 2.0-Oxford Nanopore Technologies (ONT)**

For Direct cDNA Native Barcoding Sequencing (SQK-DCS109 with EXP-NBD104, oxford nanopore technologies, Oxford, UK), 100ng of poly(A) RNA was used for the library preparation. Using H minus Reverse Transcriptase (Invitrogen, Waltham, Massachusetts, United States), complementary DNA (cDNA) was synthesised, followed by RNA degradation and second-strand synthesis of cDNA. Double-stranded cDNA was used for the end preparation, followed by native barcoding and adaptor ligation (all steps were followed, according to the manufacturer’s instructions). Ligated cDNA was loaded on the flow cell (R9) in MinION Libraries were sequenced specifying 72 hours on the ONT MinION using R9.4.1 flow cells and MinKNOW (v21.06.10, Microsoft Windows OS based) to generate FAST5 files. FAST5 files were base-called with CPU based Guppy basecaller (v.5.0.11) (ONT) to create summary text files and FASTQ files of the reads for further downstream analysis.

**Sample pre-processing and Quality assessment**

Demultiplexing of the pooled samples and adapter removal was carried out using inbuilt algorithm of Minknow. Linux Long Time Support (v.20.04) Operating System was used for all the analysis. A comprehensive report of the sequencing was generated by NanoComp (https://github.com/NanoComp/h5utils) tool and sequencing quality assessment was done by FastQc tool (https://www.bioinformatics.babraham.ac.uk/projects/fastqc/). Sample reads were subjected to a minimum phred quality score of 9 and reads with lower quality were filtered out using NanoFilt (https://github.com/wdecoster/nanofilt) tool. Some initial reads are usually prone to low base-calling quality. Hence, 50bp of each initial reads from every sample were filtered out for quality maintenance using NanoFilt tool only. All the samples were subjected to quality assessment before and after quality filtering.

**Table S1. Sequencing and mapping metrics**

Average base calling phred quality score (q) was found to be more than 10 for each sample (range 8-30). Read base quality ≥ 9 was considered for DGE analysis. N50 of read length for sequenced pooled samples was more than 900 bp (range 900bp-1.4kb). Average read-length was more than 800bp for each sample (range 200bp-4.6kb). Numbers of reads were more than 500,000 for each sample. Total number of passed (>7 q score) base sequenced for each sample was found to be more than 500 million with one sample going as much as 1 billion implying a coverage of 16X to 35X. More than 60% mapping coverage was found for each sample.

| **Sample** | **Mean read length (bp)** | **Mean read quality (q-score)** | **Read number (N)** | **Total base number (N)** | **Read length N50 (bp)** |
| --- | --- | --- | --- | --- | --- |
| Cs1 | 873.1 | 10.9 | 825,250 | 720,547,331 | 1,209 |
| Cs2 | 808.8 | 11.1 | 904,886 | 731,834,535 | 962 |
| Cs3 | 955.5 | 11 | 526,857 | 503,435,179 | 1,251 |
| Cs4 | 1045.3 | 11 | 1,066,901 | 1,115,184,646 | 1,393 |
| Mw_Cs1 | 941.5 | 10.8 | 766,585 | 721,729,757 | 1,196 |
| Mw_Cs2 | 857.8 | 10.7 | 1,053,022 | 903,322,821 | 1,134 |
| Mw_Cs3 | 881.6 | 10.7 | 580,930 | 512,171,601 | 1,109 |
| Mw_Cs4 | 817.7 | 11.1 | 846,667 | 692,33,369 | 956 |

**Differential Gene Expression analysis**

DGE was done using “pipeline-transcriptome-de” (https://github.com/nanoporetech/pipeline-transcriptome-de) pipeline. This pipeline from nanopore tech uses snakemake, minimap2, salmon, edgeR, DEXSeq and stageR to automate DGE workflows on long read data. Pipeline was set to make only reads aligned to minimum 3 samples to be considered for analysis. A separate conda (https://docs.conda.io/en/latest/#) environment was created on Linux OS to host this pipeline. Quantification and DGE was done by R language based (https://www.r-project.org/) tool edgeR (https://bioconductor.org/packages/release/bioc/html/edgeR.html) employing gene-wise negative binomial regression model and normalisation factor (Transcript Mean of M-value) for each sequence library. Differentially expressed genes (DEGs) with log2Fold Change (log2Fc) ≥0.5, ≤-0.5 and associated p-value <0.05 were selected as significant for further analysis. Annotation of DEGs was fetched from ENSEMBL database (https://asia.ensembl.org/index.html) using R based biomaRt package (https://bioconductor.org/packages/release/bioc/html/biomaRt.html). All the file compilation was ultimately done using Microsoft Excel and Linux Libre Office calc.

Hierarchical clustering analyses was performed between all four samples of each group to generate Heatmap from normalised log2 counts per million (log2CPM) expression values using R/Shiny based rnaseqDRaMA (https://hssgenomics.shinyapps.io/RNAseq_DRaMA/) package. Comparative gene expression boxplot on the basis of log2CPM expression values between Mw + ChAdOx1 and ChAdOx1 group were generated from START (https://kcvi.shinyapps.io/START/) tool. Volcano plot involving all DEGs were created using R based ggplot2 (https://ggplot2.tidyverse.org/) package.

**Gene Ontology (GO) Pathway Analysis**

Pathway enrichment analysis was done for significant DEGs using R based clusterProfiler (https://bioconductor.org/packages/release/bioc/html/clusterProfiler.html, v.4.2.1) tool employing Gene Ontology database. Pathways related to ADCC and innate immune inflammatory pathways were considered for focused analysis. An adjusted *p*-value threshold of ≤0.05 was considered for this study.

**Figure S1:**

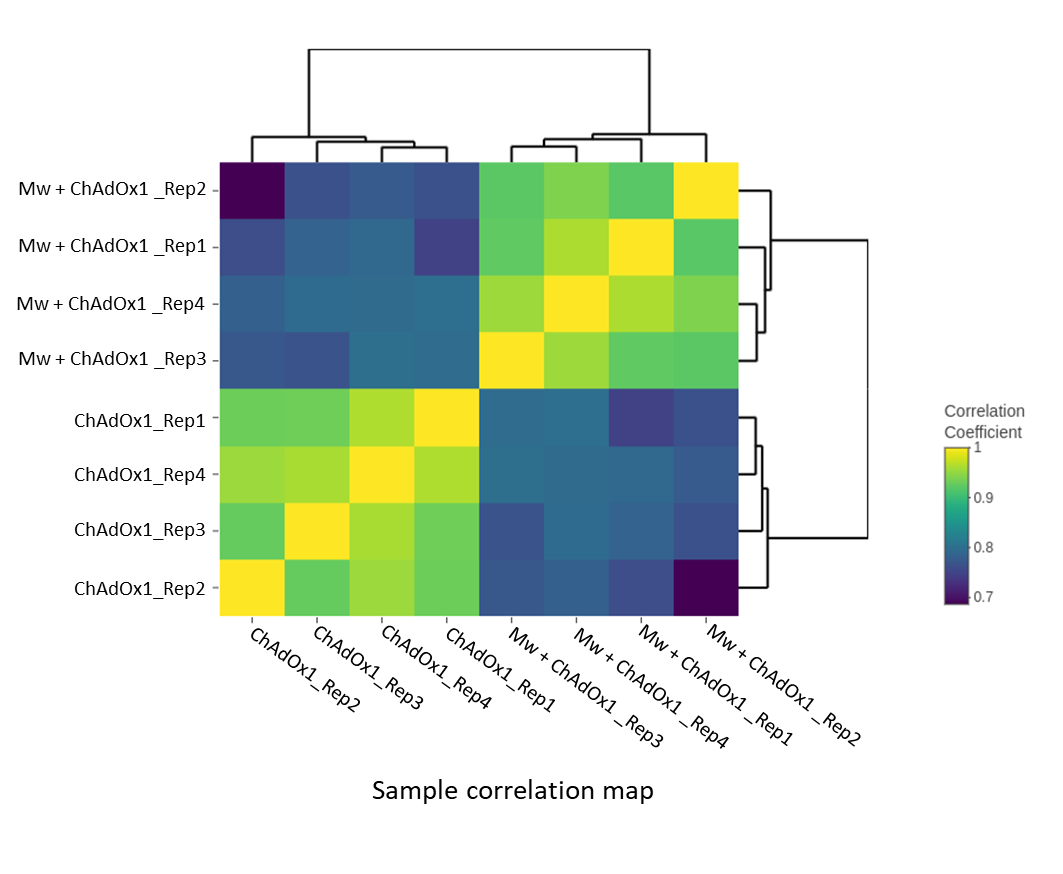

**Figure S1: Correlation matrix of 8 RNA-Seq libraries**. Pairwise Pearson correlation coefficients (PCC) were calculated for comparison among transcriptomes of Mw + ChAdOx1 and ChAdOx1 group samples. Samples were hierarchically clustered with the Euclidean distance method. The colour scale indicates the degree of correlation. The sample correlation matrix was generated using R/Shiny based rnaseqDRaMA package.

**Figure S2:**

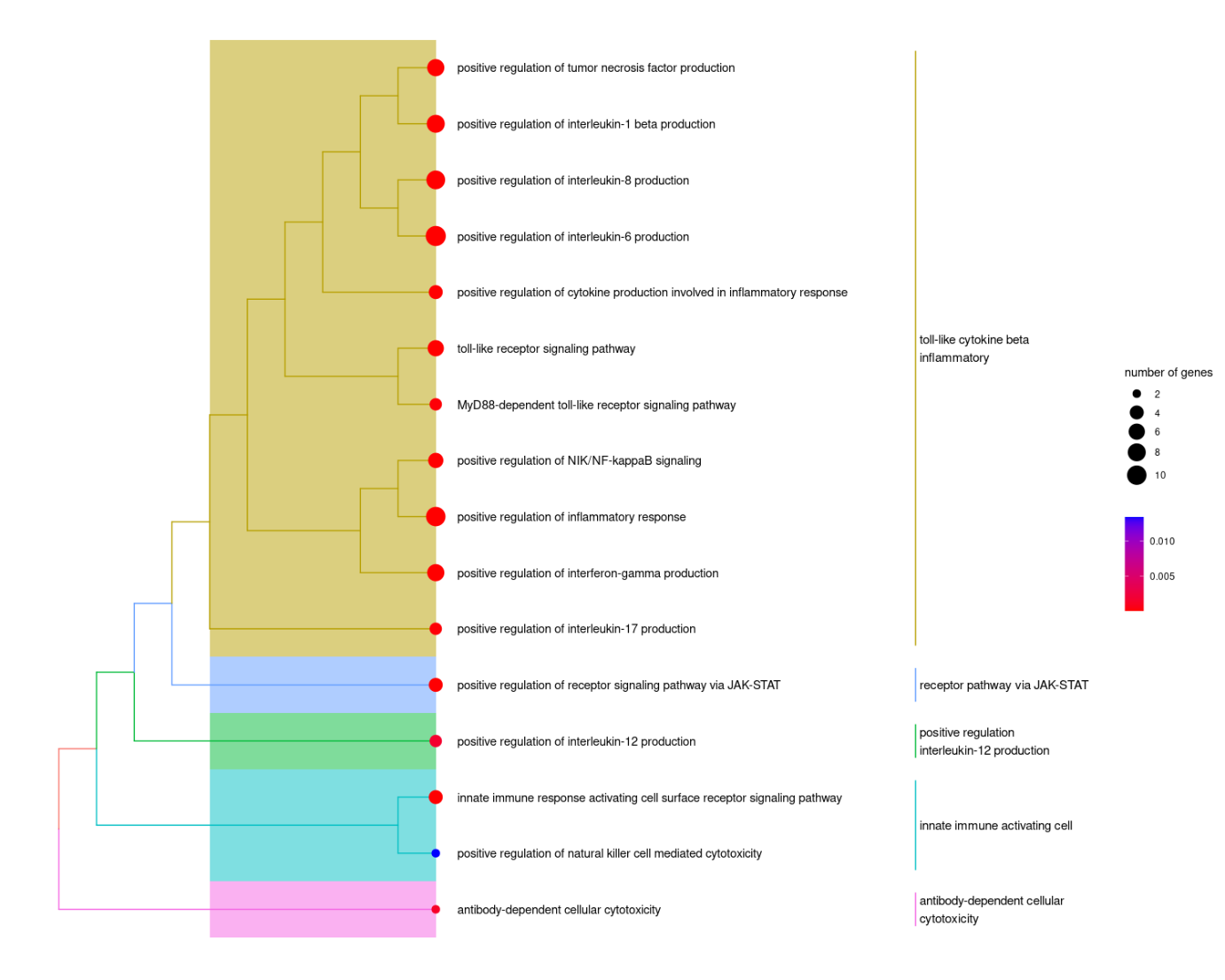

**Figure S2: Tree plot depicting clustering of enriched GO terms based on the ‘average’ hierarchical clustering method**. Subtrees are labelled using high-frequency GO words. The colour gradient scale indicates the adjusted p-value of enriched GO terms. (Fisher’s exact test using treeplot function in R based clusterProfiler package, multiple test correction by Benjamini-Hochberg method, adj. p-value <0.05)

**Analysis of the relative efficacy of Mw in prevention of COVID-19**

**Table S2. SARS-CoV-2 infections in Mw + ChAdOx1 and ChAdOx1 group**

|  | **Infected** | **Non-infected** | **Total** |
| --- | --- | --- | --- |
| **Mw + ChAdOx1 group** | 2 | 48 | 50 |
| **ChAdOx1 group** | 36 | 165 | 201 |
| **Total** | 38 | 213 | 251 |

**Table S3. Incidence rate of SARS-CoV-2 infections.**

|  | **Incidence Rate** |
| --- | --- |
| **Mw + ChAdOx1 group** | 6.7/10,000 person days |
| **ChAdOx1 group** | 29.85/10,000 person days |

**Table S4. Incidence Rate Ratio of SARS-CoV-2 infections and Mw efficacy.**

| **Parameter** | **Estimate** | ***p*-value** | **95% CI** |
| --- | --- | --- | --- |
| Incidence Rate Ratio (IRR) | 0.22 | 0.02 | 0.05 to 0.9 |
| Attack rate in ChAdOx1 group (ARU) | 0.18 |  |  |
| Attack rate in Mw + ChAdOx1 group (ARV) | 0.04 |  |  |
| Mw efficacy (%) | 77.78% |  |  |
| Absolute Risk Reduction (ARR) | 0.14 |  | 0.06 to 0.21 |
| Number Needed to Treat (NNT) | 7.2 |  | 4.7 to 15.8 |

**Table S5. Multivariate analysis of outcomes**

| Variable | Hazard Ratio (95% CI) | *p*-value |
| --- | --- | --- |
| Mw  ChAdOx1 both dose | 0.46 (0.11-1.93) | 0.008 |

**Table S6: Statistical Method for calculation of efficacy of Mw**

|  | Infected | Non-infected | Total |
| --- | --- | --- | --- |
| Mw + ChAdOx1 group | a | b | n1 |
| ChAdOx1 group | c | d | n2 |

Attack rate in ChAdOx1 group (ARU) = c/n2

Attack rate in Mw + ChAdOx1 group (ARV) = a/n1

Incidence Risk Ratio (IRR)/ Relative Risk = ARV/ARU

Absolute Risk Reduction (ARR) = ARU- ARV

Number needed to treat (NNT) =1/ARR

Vaccine Efficacy (%) = ((ARU-ARV)/ARU) *100

95% CI Vaccine efficacy (%) = 1- IRR CI
